# High Rate of SARS-CoV-2 Transmission due to Choir Practice in France at the Beginning of the COVID-19 Pandemic

**DOI:** 10.1101/2020.07.19.20145326

**Authors:** Nathalie Charlotte

## Abstract

**Background:** There has been little focus on the individual risk of acquiring COVID-19 related to choir practice.

**Methods:** We report the case of a high transmission rate of SARS-CoV-2 linked to an indoor choir rehearsal in France in March 2020 at the beginning of the COVID-19 pandemic.

**Results:** A total of 27 participants, including 25 male singers, a conductor and an accompanist attended a choir practice on March 12, 2020. The practice was indoor and took place in a non-ventilated space of 45 m^2^. No choir member reported having been symptomatic for COVID-19 between March 2 and March 12.The mean age of the participants was 66.9 (range 35-86) years. 70% of the participants (19 of 27) were diagnosed with COVID-19 from 1 to 12 days after the rehearsal with a median of 5.1 days. 36% of the cases needed a hospitalization (7/19), and 21% (4/19) were admitted to an ICU. The index cases were possibly multiple.

**Discussion:** The choir practice was planned in March 2020 at a period when the number of new cases of COVID-19 began to grow exponentially in France because SARS-CoV-2 was actively circulating. The secondary attack rate (70%) was much higher than it is described within households (10-20%) and among close contacts made outside households (0-5%). Singing might have contributed to enhance SARS-CoV-2 person-to-person transmission through emission of droplets and aerosolization in a closed non ventilated space with a relative high number of people including multiple pre-symptomatic suspected index cases.

**Conclusion:** Indoor choir practice should be suspended during SARS-CoV-2 outbreaks. Further studies are necessary to test the spread of the virus by the act of singing. As the benefits of the barrier measures and social distancing are known to be effective in terms of a reduction in the incidence of the COVID-19, experts’ recommendations concerning the resuming of choir practice are necessary.

## Introduction

World Health Organization (WHO) declared the COVID-19 outbreak a pandemic on March 11, 2020^**1**^. In response, all countries have implemented large scale public health and social measures in an attempt to reduce the transmission of SARS-CoV-2 and minimize the impact of the epidemic. Some social activities as fitness practice ^**2**^, night-clubbing ^**3**^, Zumba practice ^**2**^, or working in open space offices^**4**^ have been described as super spreader events for COVID-19. Approximatively 4.5% of the European population actively participate in collective singing activities^**5**^. There has been very little focus on the individual risk of acquiring COVID-19 related to choir practice. We report the case of a high transmission rate of SARS-CoV-2 linked to an indoor choir rehearsal in France at the beginning of the pandemic.

## Methods – case analysis

The data were obtained from the participants via the collection of anonymous responses to a questionnaire. The participants were the persons who attended the March 12, 2020 choir practice i.e. the choristers, the conductor and the accompanist. The president of the choir collected the questionnaires filled by the participants. The response rate to the questionnaire was 100%. The president and the conductor of the choir were interviewed by telephone. The President of a second choir designated by choir number 2 was also interviewed by telephone. The COVID-19 diagnosis were established by the general practitioners of the attendees or by hospital physicians for those who were hospitalized for life threatening COVID-19.

On March 12, 2020, 27 men (25 singers, 1 accompanist, 1 conductor) attended a choir 2-hours singing practice in France. The mean age of the participants was 66.9 (range 35-86) years. It took place inside a 45 m^2^ and 3 m high room (mean surface available per person of 1.6 m^2^). The room wasn’t ventilated. The members were sitting less close to one another than usual, but at a distance of less than 6 feet. Socialization acts were avoided during the practice (no shaking hands, no food sharing and no pauses). The male choir had a previous rehearsal on March 2. No choir member reported having been symptomatic for COVID-19 between March 2 and March 12. The individuals were split into 2 groups of 11 tenors and 14 basses. The mean age of the tenors was 66.9 (range 35-86) years. The mean age of the basses was 69.1 (range 48-84) years. The conductor was standing in front of the choir, the accompanist was near the tenors. (Figure 1).

**Figure 1:**
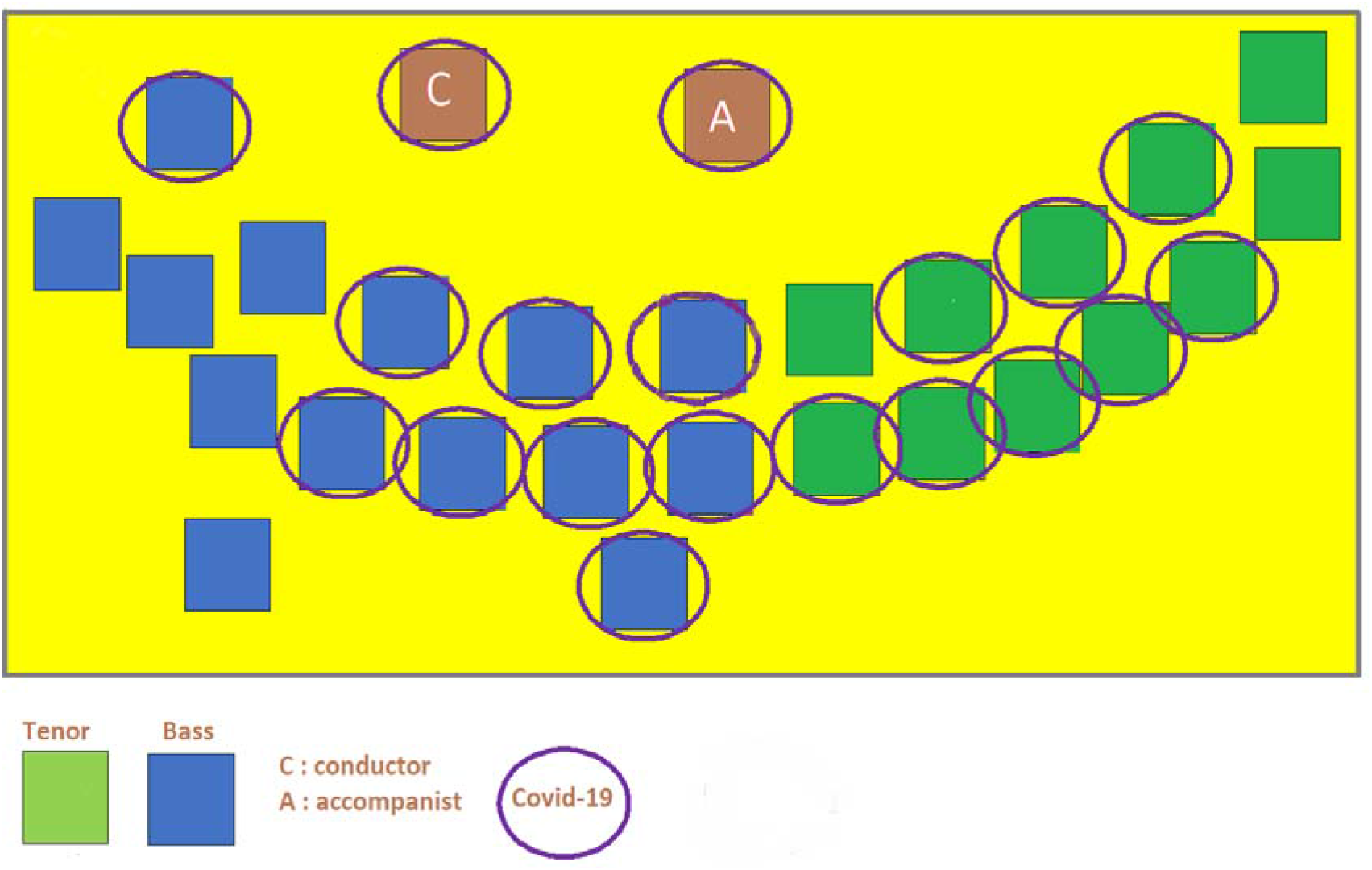
Sketch showing the position of the attendees in the practice room on May 12, 2020.

Nineteen COVID-19 cases were identified among the members of the ensemble (Figure 2). A confirmed case was defined as an individual who has tested positive for SARS-CoV-2 by RT-PCR test performed on a nasopharyngeal sample and/or a case presenting a severe COVID-19 requiring hospitalization. A probable case was defined as a person presenting an acute respiratory illness relating to COVID-19 by a general practitioner 1 to 14 days following the rehearsal without having been evaluated with an RT-PCR test. Seven confirmed cases and 12 probable cases were identified. The dates of onset of symptoms among the cases ranged from 13/03 to 22/03 i.e. 1 to 10 days after the practice (median = 5 days, range 1-13 days). The secondary attack rate (SAR) was 33% among the confirmed cases and the overall attack rate was 70%. The SAR was 72% (8/11) among the tenors, of 64% (9/14) among the basses, and of 100% (2/2) among the non-choristers (conductor and accompanist) who were standing in front of the singers. The index cases were possibly multiple. Of 2 attendees who had a close contact with a COVID-19 case within 7 days before the practice, 1 develop COVID-19 symptoms 4 days after the rehearsal. One chorister (tenor) who tested positive for COVID-19 has had close contacts with the members of another choir (choir number 2) during 2 practices in a nearby area on March 9 and March 11. Several of the choristers of choir number 2 began to report COVID-19 symptoms on March 14 and the following dates.

**Figure 2:**
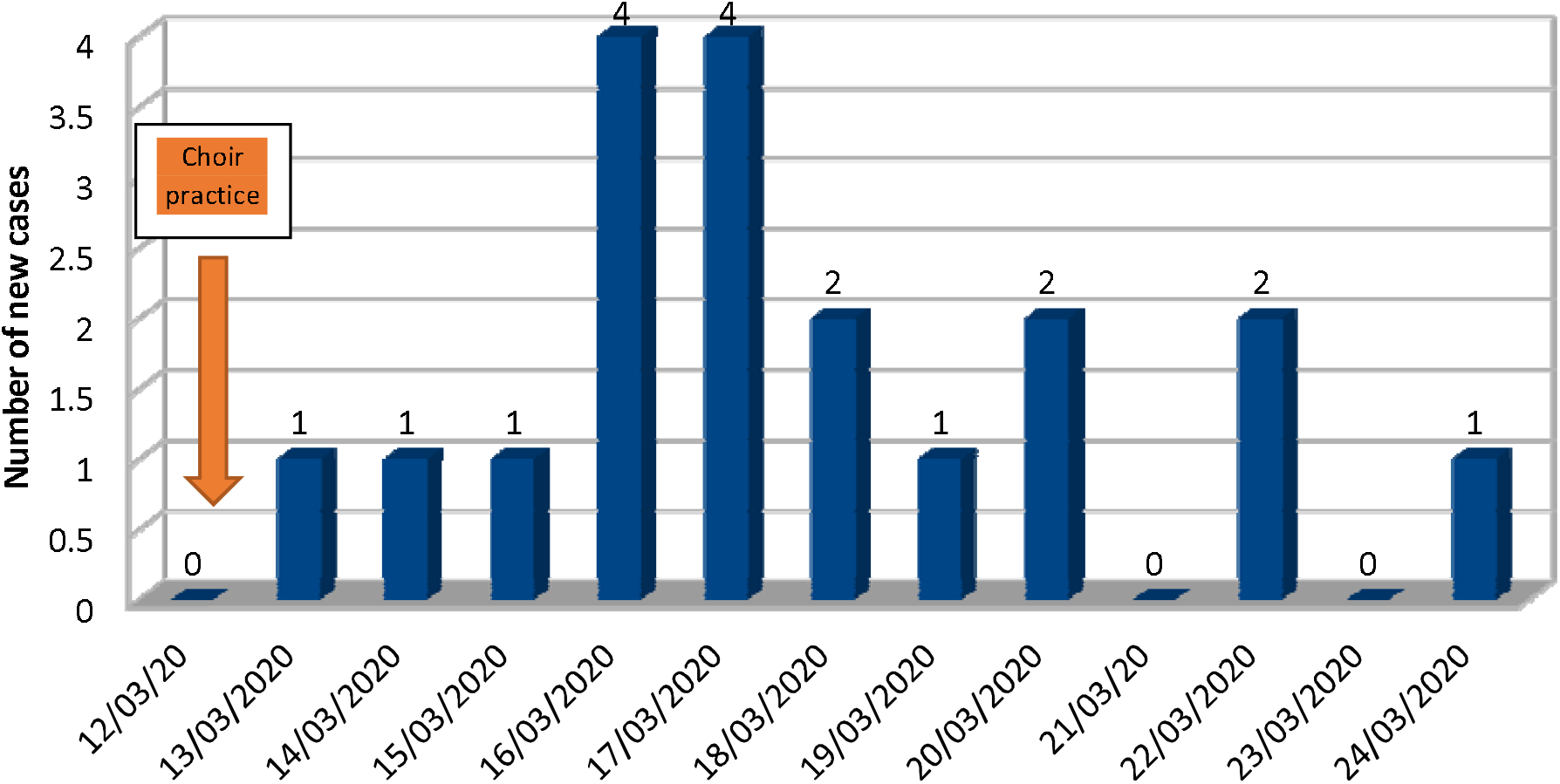
Incidence of secondary COVID-19 cases among the choir attendees.

The mean age of the individuals who developed a severe COVID-19 defined by the need for a hospitalization, was 70.5 years (range 53-78). Seven of the 19 persons who became ill were hospitalized (36%), whose 57% (4/7) which represents 21% (4/19) of all patients were treated in an intensive care unit. The mean age of those who developed a mild COVID-19 (64%) was 60.9 years, (range 35-77). The fatality rate was 0%. Thirteen tertiary cases were later reported, mostly household contacts.

## Discussion

From end February to end March 2020, SARS-CoV-2 was actively circulating in Europe. Several clusters of COVID-19 cases linked to choir practice were reported by the general media. In Whir au Val (Haut-Rhin, France), after a rehearsal of the Chorale de Saint Martin on February 28 with 20 choristers, 69 secondary cases were later identified and 9 persons died^**6**^. The Amsterdam Gemengd Koor (130 choristers) gave a 3-hour concert on March 8 at the Concertgebouw of Amsterdam (The Netherlands). The occurrence of 102 secondary COVID-19 cases was reported, including 4 fatal cases^**7**^. In Germany, within 2 weeks after gathering for a rehearsal on March 9, thirty members of the Berlin Cathedral Choir’s 80 singers tested positive for COVID-19, and a further 30 reported COVID-19 symptoms^**8**^.

In the USA, Skagit County Public Health (Washington) investigated a cluster of 53 persons among 61 choristers after a choir practice on March 10. Two of them died of COVID-19^**9**^.

In France, the number of new cases of COVID-19 began to grow exponentially in March^**10**^. In the absence of effective treatment for the disease, efforts to control COVID-19 pandemic have relied on personal preventive actions, physical distancing, stay-at-home orders, school closures, and workplace restrictions adopted at the national level. On March 9 the gathering of more than 1 000 people was forbidden. On March 12 the closure of all schools and universities was ordered. On March 13 those restrictions were extended to the gathering of more than 100 people. The stay-at-home order was effective on March 17 after more than a thousand new confirmed cases were reported within 24 hours ^**10**^. On the date the rehearsal took place, people in France were not yet familiar with the barrier measures.

The incubation period of SARS-CoV-2 is known to be 1 to 14 days with a median of 5.1 days^**11**^. The March 12 practice is the likely point-source exposure event because the surge of COVID-19 cases occurred 5 days later. The individual whose onset of symptoms was March 13 (within 24 hours after the practice), might have been a pre-symptomatic or asymptomatic carrier of SARS-CoV-2 at the time of the rehearsal, which could explain the shortness of the delay between the date of the event and the onset of the illness^**12**^. As all the choristers were asymptomatic at the time of the practice, we can only presume that the three attendees who declared having had close contact with a COVID-19 case within 7 days before the event may have been index cases. Evidence of transmission of SARS-CoV-2 from asymptomatic or pre-symptomatic individuals has been well established by several authors^**12**, **13**, **14**^.

The basic reproduction number of SARS-CoV-2 is approximately of 2.2 to 2.5^**15**^. The estimated secondary attack rate of the virus (SAR) within households is 10–20%, with a much smaller SAR among close contacts made outside households, (0–5%) ^**16**^. In comparison, we found a much higher SAR which shows that SARS-CoV-2 has had the potential to spread widely during the choir rehearsal under its peculiar environmental and circumstances: indoor setting, narrow and non-ventilated space, shortened inter-individual distances. A recent study shows that the transmission of SARS-CoV-2 is likely at much higher risk in enclosed spaces than outside^**17**^. In addition, some other high human density environments like workplaces have been reported as important risk sites for the spread of SARS-CoV-2 and potential sources of further transmission^**4**^. For that reason, at the on-going re-opening phase of the pandemic, a minimum surface area of 4 m^2^ available per person is recommended by the French Work Ministry for people working in the same room.^**18**^

SARS-CoV-2 is transmitted between people by infectious droplets emitted in close proximity to the nose, mouth or eyes of a susceptible person^**19**^. A recent study emphasizes the role of airborne transmission for the spread of the virus^**20**^. Smaller droplets are able to be propelled further than 3 to 6 feet and to remain airborne longer after certain respiratory emissions^**21**, **22**^, particularly in poorly ventilated spaces^**23**^. Singing might have contributed to enhance SARS-CoV-2 person-to-person transmission through emission of droplets and aerosolization in a closed space with a relative high number of individuals. WHO recently acknowledged that transmission of the virus by aerosols may have been responsible for reported outbreaks of COVID-19 in some closed settings, such as places of worship or places of work where people may be shouting, talking, or singing^**24**^? We can also assume that if the asymptomatic index cases were multiple, they might have behaved as a single super-spreader case^**25**^ and exposed the participants to a high viral load during the activity of singing. Furthermore, despite avoiding social relations during the practice, it might have been occasions of facilitating the transmission of the virus like speaking face to face with one another shortly but recurrently.

This investigation has a number of limitations. The choristers were not interviewed by the author. The clinical features of the COVID-19 presentation of the cases were not available, prohibiting a detailed description of symptoms. The index case-patients were not identified. A majority of patients were not tested for SARS-CoV-2, which might have led to under-ascertainment of infections, particularly for those who were pre-symptomatic or asymptomatic. At mid-March 2020, France was at the third stage of the epidemic with the SARS-CoV-2 virus actively circulating countrywide. Testing patients reporting mild symptoms wasn’t recommended in this context by the French High Council for Public Health (Haut Conseil de la Santé Publique, HCSP) who prioritized the access to diagnostic tests to patients at risk of severe forms of Covid-19 and to healthcare workers^**26**^.

## Conclusion

At the end of winter 2020 in Europe, choir practice indoors was a super spreader event because of the active circulation of the virus in the population at this moment, the characteristics of the viral transmission via close human contacts probably enhanced by the act of singing, the social gathering and the potential presence of multiple index cases spreading the virus. Indoor choir practice should be suspended during SARS-CoV-2 outbreaks. Further studies are necessary to test the spread of the virus by the act of singing. As the benefits of the barrier measures and social distancing are known to be effective in terms of a reduction in the incidence of the COVID-19, the question of the resuming of choir practice is being raised: experts’ recommendations are necessary and must be set out to provide operational guidelines to singers at an amateur as well as a professional level.

## Data Availability

The data are available on demand to the author.

## Acknowledgements

We sincerely thank the president of the choir and its conductor for their precious participation in this investigation.

